# Performance and usability evaluation of three LDH-based malaria rapid diagnostic tests in Kédougou, Senegal

**DOI:** 10.1101/2024.12.12.24318945

**Authors:** Babacar Souleymane Sambe, Stephanie Zobrist, William Sheahan, Divya Soni, Aissatou Diagne, Ibrahima Sarr, Arona Sabene Diatta, Serigne Ousmane Mbacke Diaw, Allison Golden, Hannah Slater, Ihn Kyung Jang, Nerie Roa, Sampa Pal, Fatoumata Diene Sarr, Joseph Faye, Inès Vigan-Womas, Yakou Dieye, Moustapha Cisse, Gonzalo J Domingo, Makhtar Niang

**Affiliations:** Institut Pasteur de Dakar, Dakar, Senegal; PATH, Seattle, United States; PATH, New Delhi, India; PATH, Dakar, Senegal

**Keywords:** Malaria, diagnosis, rapid diagnostic test, lactate dehydrogenase, histidine rich protein 2

## Abstract

**Background:** The emergence of *pfhrp2/3*-deleted parasites threatens histidine-rich protein 2 (HRP2)-based malaria rapid diagnostic test (RDT) performance. RDTs targeting *Plasmodium falciparum* (*Pf*) lactate dehydrogenase (LDH) may address current product limitations and improve case management.

**Objectives:** To evaluate the performance and usability of three LDH-based RDTs in febrile patients.

**Methods:** A cross-sectional diagnostic accuracy study was conducted in Kédougou, Senegal. Capillary blood was tested using the SD Bioline Ag Pf (#05FK50) and three LDH-based RDTs: BIOCREDIT Malaria Ag Pf (pLDH), BIOCREDIT Pf (pLDH/HRPII), and BIOCREDIT Pf/Pv (pLDH/pLDH) (Rapigen Inc., Republic of Korea). Venous blood was collected to repeat the BIOCREDIT RDTs and conduct microscopy. Frozen venous specimens were tested using a reference PCR assay. A quantitative multiplex malaria antigen assay measured antigen concentration. RDT performance was determined and analyzed as a function of antigen concentration distribution. Usability of the *Pf*-only BIOCREDIT tests was evaluated using a questionnaire.

**Results:** 154/220 participants (70%) were *Pf*-positive by PCR. The Pf (pLDH/HRPII) test demonstrated the highest sensitivity at 78% (70.9%–84.5%); specificity was 89% (79.4%–95.6%). All RDTs performed better than microscopy (53% sensitivity). RDTs performed better when compared to antigen concentration over PCR results. Improved sensitivity of the Pf (pLDH/HRPII) test was driven by the HRP2 line. Line intensity correlated with antigen concentration. Predicted sensitivity using the analytical limit of detection (LOD) was comparable to observed sensitivity. RDTs demonstrated acceptable usability.

**Conclusions:** Both HRP2 and LDH contributed to the sensitivity of the best-performing *Pf-*RDT. RDT analytical LODs can be used to predict performance in populations with known antigen concentrations.

## Introduction

Accurate and timely diagnosis of malaria is essential to ensure effective treatment for patients and accelerate control and elimination efforts. Lateral flow immunochromatographic rapid diagnostic tests (RDTs) for malaria have been widely accepted in endemic settings due to their simplicity, low cost, minimal infrastructure requirements, and rapid time to result.^1,2^

Malaria RDTs function by detecting specific protein antigens expressed by the malaria parasite in the blood of infected individuals.^3^ Histidine-rich protein 2 (HRP2) and lactate dehydrogenase (LDH) are two common antigens targeted by malaria RDTs. The vast majority of RDTs used for the diagnosis of *Plasmodium falciparum* (*Pf*) malaria target HRP2, as this antigen is specific to *Pf* and cannot be produced by other malarial species.^4^ To date, HRP2 has been the preferred target for *Pf* RDTs due to its abundant production by the parasite, as well as greater heat stability and clinical sensitivity as compared to PfLDH-based RDTs.^5–11^ Senegal, along with many other African countries where *Pf* is the predominant species,^12,13^ currently relies primarily on HRP2-based RDTs for malaria diagnosis.

There are significant limitations to the widespread use of HRP2-based *Pf* RDTs. The HRP2 antigen can persist in the peripheral bloodstream for multiple weeks following parasite clearance and can, therefore, result in false-positive results among individuals who have recently received treatment. Most importantly, the performance of HRP2-based RDTs is threatened by increasing reports of parasites with deleted *hrp2/hrp3* genes.^14,15^ In settings with a significant prevalence of *hrp2/hrp3* gene deletions in symptomatic populations, the World Health Organization (WHO) advises switching to RDTs that do not rely exclusively on HRP2 detection.^16^ Several studies have documented observations of *Pf hrp2/hrp3* gene deletions in Senegal.^17,18^ Recent modeling suggests that deletions identified in Western Africa are projected to increase,^19^ and, as such, further monitoring of deletion prevalence and impact on RDT performance in this setting is warranted. A switch to PfLDH-based tests must take into account the significantly higher limits of detection (LOD) for LDH, and consequently the lower clinical sensitivity of these tests.^20^ For this reason, improvements in the performance of PfLDH-based tests are needed in order to minimize tradeoffs in sensitivity.

Rapigen Inc. (Republic of Korea) has developed three novel, LDH-based malaria RDTs:

- The BIOCREDIT Malaria Ag Pf (pLDH/HRPII) RDT, with two test lines for HRP2 and PfLDH
- The BIOCREDIT Malaria Ag Pf/Pv (pLDH/pLDH) RDT, with two test lines for PfLDH and PvLDH
- The BIOCREDIT Malaria Ag Pf (pLDH) RDT, with one test line for PfLDH.

Analytical benchmarking and clinical sensitivity modeling for these tests indicate that they have improved LODs for LDH that may result in higher clinical performance in terms of sensitivity toward *Pf* infections with *hrp2/hrp3* gene deletions and for *Plasmodium vivax (Pv)* clinical infections.^20^ Additionally, previous evaluations of these tests in Ghana, Burundi, Uganda, Djibouti, and the Republic of Korea have also demonstrated promising clinical performance.^21–25^

This study aimed to evaluate the clinical performance of these three malaria RDTs among a febrile population in Kédougou, Senegal, in comparison to a reference polymerase chain reaction (PCR) assay and antigen concentration quantification. The performance of microscopy and a currently available HRP2-based comparator RDT were also evaluated in the same population in order to enable informed decision-making regarding the recommendation of new, highly sensitive point-of-care tools for malaria.

## Materials and methods

### Ethical approval

This study was reviewed and approved by the Comité National d’Ethique pour la Recherche en Santé (CNERS) [00000126/MSAS/CNERS/SP], Sénégal, and WIRB-Copernicus Group (WCG) [1313427] Institutional Review Boards (IRBs). Written informed consent was obtained for all study participants. For minors, consent was provided by legal guardians, and children over the age of 7 also provided assent.

### Study design and population

Between November 2021 and February 2022, a cross-sectional diagnostic accuracy study of patients presenting with febrile symptoms was conducted in Kédougou, Senegal. The Kédougou region borders Guinea and Mali and experiences a high burden of *Pf* malaria with moderate seasonality.^26,27^ Patients aged 6 months and older presenting with febrile symptoms were recruited from five health facilities: the Kédougou Health Center, and the Tomboronkoto, Dalaba, Bandafassi, and Bantako health posts.

Individuals weighing less than 8 kg or those who had a serious illness, as determined by the health care provider, were excluded from participation. Given the high malaria transmission and parasite prevalence in the study location, it was expected that some participants would either become reinfected or continue to exhibit malaria during the study period and return to the health center/post. In such cases, the individual’s status as a returning participant was recorded, but all testing was repeated, and they were treated in the study analysis as a unique sample.

### Study procedures and RDT testing

Following enrollment, participants completed a brief questionnaire to collect information on demographics, current health status, and medical history. Next, trained study staff collected capillary blood samples, performed the standard-of-care malaria RDT (the SD BIOLINE Malaria Ag P.f [#05FK50]), and conducted the three investigational tests: the Rapigen BIOCREDIT Malaria Ag Pf (pLDH/HRPII), BIOCREDIT Malaria Ag Pf (pLDH), and BIOCREDIT Malaria Ag Pf/Pv (pLDH/pLDH). All tests were conducted according to the manufacturer’s instructions and read within the specified timeframes. Test and control line intensities of the Rapigen test results were assigned based on comparison of test and control line observed intensities to 16-point (test) and 5-point (control) intensity scales on a card provided by Rapigen. All invalid test results were recorded. Results from the standard-of-care RDT were used to inform clinical care, and all patients found positive were treated according to national guidelines.

Next, 4 mL of venous blood was collected into an EDTA tube and transported under cold chain to the Institut Pasteur field station in Kédougou. At the field station, the venous blood was aliquoted and the three BIOCREDIT RDTs were repeated. All test operators at the field station were blinded to the clinic RDT results. One aliquot of venous blood was also used to prepare microscopy slides, and the remaining aliquots were frozen at -20°C. Microscopy slides were read by a trained, blinded microscopist at the Institut Pasteur de Dakar laboratory in Dakar, Senegal. Frozen venous blood specimens were shipped on ice to PATH (Seattle, USA) for reference PCR testing and confirmatory antigen concentration determination. Operators performing the reference and confirmatory assays were blinded to all RDT and microscopy results and vice versa.

### Reference PCR testing

Frozen whole blood in EDTA was analyzed for the detection of *Pf* nucleic acid using a real-time PCR assay. A photoinduced electron transfer (PET) PCR assay for P*f* was conducted at PATH. DNA was extracted from 100 µL of frozen whole blood using QIAamp DNA Mini Kits (QUIAGEN, Valencia, CA, cat #51106) and eluted into 100 µL of elution buffer and stored at -20°C. All samples were screened for P*f* using the forward (5’-ACC CCT CGC CTG GTG TTT TT-3’) and reverse (HEX-5’-agg cgg ata ccg cct ggT CGG GCC CCA AAA ATA GGA A-3’) self-quenching primers, as established by the Centers for Disease Control and Prevention.^28^ The reaction was performed in a 20 μL reaction containing 2X TaqMan Environmental buffer 2.0 (Applied Biosystems, Grand Island, NY, Cat # 4398044), 62.5 nM each of forward and reverse primers and 5 μL of template DNA. Cycling parameters were 95°C for 10 minutes, followed by 45 cycles of denaturation at 95°C for 10 seconds and annealing at 60°C for 40 seconds. Cycle threshold (Ct) values, defined as the number of cycles required for the fluorescent signal to cross the threshold (i.e., exceeds the background level), were recorded after the end of each annealing step.

Specimens with Ct values <40 were considered positive. *Pf* 3D7 with known parasitemia was used as a positive control. All assays were performed using Agilent Mx3005pro thermocyclers (Agilent Technologies, Santa Clara, CA).

### Antigen concentration determination

Antigen concentration was determined with a quantitative antigen assay: Q-Plex™ Human Malaria Array (5-Plex) (Quansys Biosciences, Logan UT, USA). This is a chemiluminescent ELISA that allows the identification of malaria infection and the simultaneous detection of human C-reactive protein (CRP) in human blood, serum, and plasma. This assay uses a quantitative sandwich enzyme immunoassay to measure malaria HRP2, PfLDH, PvLDH, PanLDH antigens, and human CRP in less than 4 hours.^29,30^ The cutoffs used for antigen positivity were as described previously.^29^

### Usability study

Two usability studies were conducted to evaluate the *Pf-*only BIOCREDIT RDTs: the BIOCREDIT Malaria Ag Pf (pLDH) and the BIOCREDIT Malaria Ag Pf (pLDH/HRPII) RDTs. Participants included health care workers, who are representative of intended end users of malaria RDTs in rural and urban settings in Senegal. Each participant evaluated only one of the two RDTs and completed a questionnaire assessing label and packaging comprehension as well as results interpretation of images of contrived RDTs.^31^

### Data management and statistical analysis

All study results were recorded on data collection forms and double-entered into REDCap® (Research Electronic Data Capture, version 12.3.3), a web-based software platform with built-in validation rules to minimize data entry errors.^32^

For the estimation of sample size, a 95% confidence interval was assumed, with an absolute precision of 0.025. In the absence of preliminary data, a 50% prevalence of *Pf* infection was assumed, yielding a target sample size of 241 participants.

Descriptive statistical analysis, including calculating point estimates, distributions, and frequencies of responses, was conducted to summarize and characterize the study population. The number and percentage of participants infected with malaria, as determined by all assays, were assessed. Diagnostic accuracy was determined by calculating the sensitivity, specificity, positive predictive value (PPV), and negative predictive value (NPV) of all RDTs and microscopy in comparison to (1) the reference PCR assay and (2) the quantitative antigen assay. The diagnostic accuracy of the quantitative antigen assay relative to the PCR reference method was also calculated. For comparisons using antigen concentrations as the reference method, the cognate target (PfLDH, HRP2, or both) was used to determine true positivity or negativity. All diagnostic performance results were reported with 95% confidence intervals (CIs) calculated using the conservative exact binomial method.

RDT performance was analyzed as a function of antigen concentration distribution and parasite density. This analysis was conducted only for the BIOCREDIT Malaria Ag Pf (pLDH/HRPII) RDT as it was the only test in this study with both PfLDH and HRP2 lines and had comparable PfLDH results to the other two BIOCREDIT products. Firstly, the relationships between HRP2 and PfLDH concentrations and parasite density, stratified by RDT result, are described using boxplots with jittered datapoints overlaid. Next, scatterplots of HRP2 versus PfLDH concentration stratified by RDT status and PCR status are used to explore visually explore the data. A sigmoidal dose response statistical model was then fitted within a Bayesian framework to estimate the relationship between antigen concentration and operator-assigned RDT line intensity scores for each antigen. Finally, a logistic regression model was fitted within a Bayesian framework to estimate the relationship between antigen concentration and the probability of RDT positivity for each antigen.^20^ In both models, antigen concentrations were log10 transformed to ensure appropriate scaling. Uninformative priors were used in the line intensity model. Both models were run for 10,000 iterations and convergence was assessed via visual checks of the coefficient trace plots.

^20^The Rapigen BIOCREDIT Pf (pLDH/HRPII) test has previously been evaluated for analytical performance against antigen concentration using a standardized laboratory panel.^20^ Golden *et al.* (2024) determined the test’s antigen LOD, defined as the concentration at which the test was expected to be positive 90% of the time, to be 525 pg/mL for HRP2 and 1,318 pg/mL for LDH. Here, we applied these cutoffs to predict the test result for each sample as detectable or not detectable by the RDT, depending on whether the measured cognate antigen concentration was above or below the LOD. The predicted clinical performance of the BIOCREDIT Malaria Ag Pf (pLDH/HRPII) RDT against both the reference PCR and antigen concentration results in this population was then calculated.

Statistical analysis was conducted using R version 4.2.1 (R Foundation for Statistical Computing, Vienna, Austria).

Usability assessment included both multiple-choice and open-ended questions, and responses were analyzed using descriptive statistics.

## Results

### Characteristics of the study population

A total of 236 participants were included in the study. Of these, 220 had venous blood available for reference assay analysis and were therefore included in the analytical sample. Of these 220 participants, capillary results for the investigational tests were not available for two participants, and one result from the BIOCREDIT Malaria Ag Pf (pLDH) test was excluded from analysis due to a data recording error. Microscopy results were available for183 participants, and a subset of specimens from 200 participants with sufficient specimen volume underwent antigen concentration testing.

Table 1 presents the demographic information of the study participants. The majority of participants (60%) were over 16 years of age, and there was an even distribution of male (49%) and female (51%) participants. The study population reported a diverse range of symptoms, and 41% were febrile at enrollment, defined as having a body temperature of ≥37.5°C.

**Table 1.** Demographic information of study participants.

|  | Value | N | Proportion |
| --- | --- | --- | --- |
| **Age (years)** |  |  |  |
| Mean (SD) | 20.42 (14.81) | 220 | - |
| Range | 0 – 71 | 220 | - |
| **Age categories** |  |  |  |
| 0-2 years | 15 | 220 | 0.07 |
| 3-5 years | 17 | 220 | 0.08 |
| 6-12 years | 41 | 220 | 0.19 |
| 13-15 years | 13 | 220 | 0.06 |
| 16-64 years | 130 | 220 | 0.59 |
| 65+ years | 4 | 220 | 0.02 |
| **Sex** |  |  |  |
| Male | 107 | 220 | 0.49 |
| Female | 113 | 220 | 0.51 |
| **Pregnancy status** |  |  |  |
| Pregnant | 9 | 113 | 0.08 |
| Not pregnant | 95 | 113 | 0.84 |
| Rather not say | 9 | 113 | 0.08 |
| **Any antimalarial drug received in the last four weeks** |  |  |  |
| Yes | 55 | 219 | 0.25 |
| No | 164 | 219 | 0.75 |
| **Symptom status** |  |  |  |
| Abdominal pain | 50 | 220 | 0.23 |
| Chills | 54 | 220 | 0.25 |
| Conjunctival redness | 2 | 220 | 0.01 |
| Cough | 43 | 220 | 0.20 |
| Dizziness/vertigo | 49 | 220 | 0.22 |
| Dysuria | 1 | 220 | 0.00 |
| Fatigue | 131 | 220 | 0.60 |
| Fever (self-report) | 173 | 220 | 0.79 |
| Headache | 151 | 220 | 0.69 |
| Nausea | 36 | 220 | 0.16 |
| Stomachache | 59 | 220 | 0.27 |
| Sweats | 13 | 220 | 0.06 |
| Vomiting | 72 | 220 | 0.33 |
| Other | 52 | 220 | 0.24 |
| **Febrile at enrollment (temperature $\geq 37.5^{\circ}\text{C}$)** |  |  |  |
| Yes | 89 | 219 | 0.41 |
| No | 130 | 219 | 0.59 |

SD, standard deviation.

Seventy percent (154/220) of enrolled participants were *Pf*-positive by reference PCR. For these PCR-confirmed *Pf-*positive cases, the mean parasite density was 729,386.5 parasites/µL, the mean HRP2 concentration was 3,399.3 ng/mL, and the mean PfLDH concentration was 556.6 ng/mL. No *Pv-*positive specimens were observed on any of the assays. One suspected *hrp2/hrp3* deletion case was identified by the criteria of HRP2-negative and PfLDH-positive antigen concentration results in this population. No invalid results were obtained on any RDTs in this study.

### Diagnostic performance of investigational and comparator RDTs against reference PCR

Table 2 presents a summary of the diagnostic performance of all investigational and comparator tests compared to the PCR reference assay by specimen type. When using the PCR reference method, microscopy showed the lowest overall performance with a sensitivity of 53% (95% CI: 44.5–62.2). The BIOCREDIT Malaria Ag Pf (pLDH/HRPII) test showed the highest sensitivity of all evaluated tests at 78% (95% CI: 70.9 – 84.5). The corresponding PPV and NPV were 94% (95% CI: 88.9–97.7) and 64% (95% CI: 53.5–73.9), respectively. The two BIOCREDIT PfLDH-only tests in this study showed the lowest performance. Both of these tests, as well as the PfLDH line alone on the BIOCREDIT Malaria Ag Pf (pLDH/HRPII) test, had lower sensitivity than the comparator HRP2-based RDT (71%; 95% CI: 63.6– 78.4). The BIOCREDIT Malaria Ag Pf (pLDH/HRPII) test also had the lowest specificity at 89% (95% CI: 79.4–95.6), with the HRP2 line showing the lowest specificity.

**Table 2.**
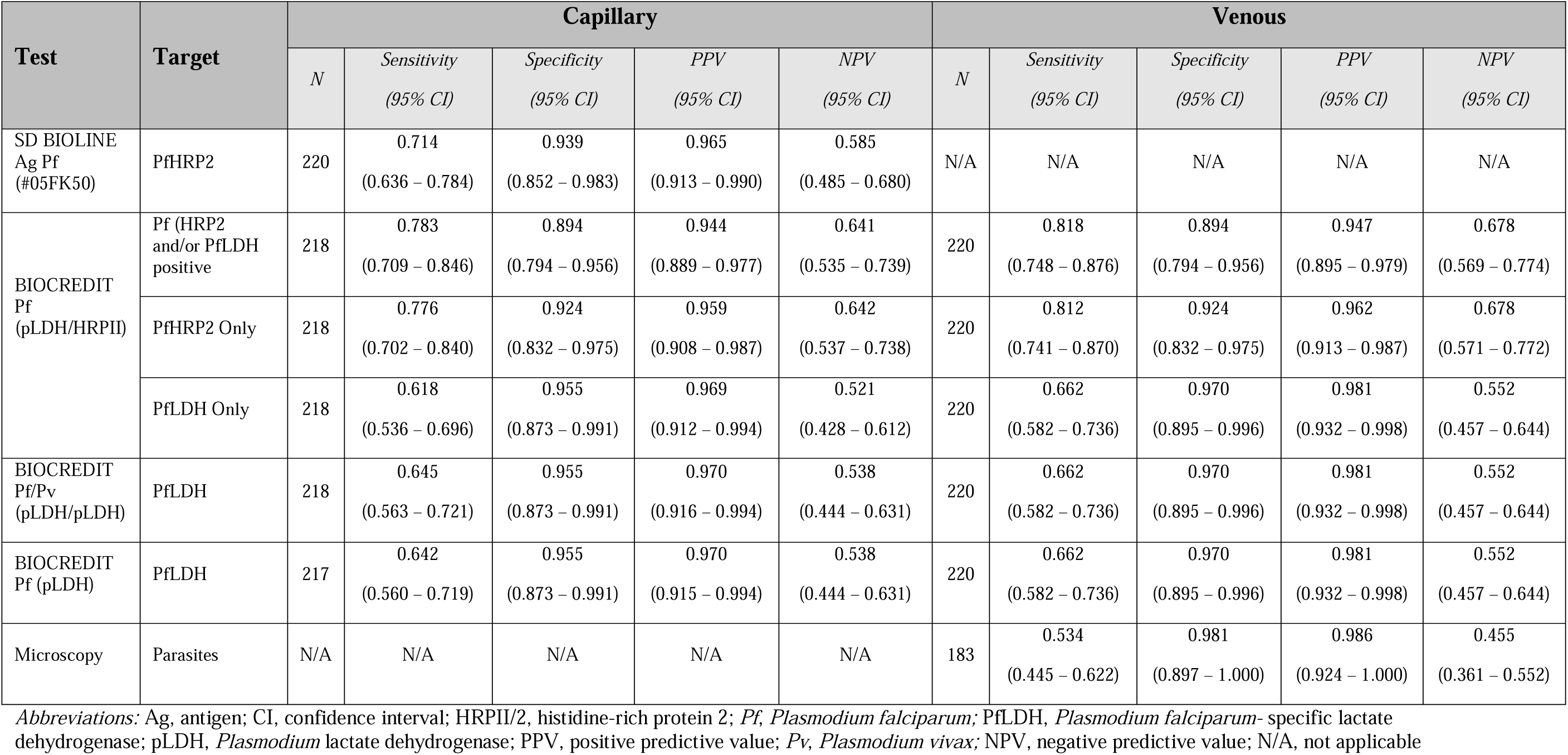
Diagnostic performance of investigational and comparator tests against reference PCR for detection of *P. falciparum* by specimen type.

| Test | Target | Capillary |  |  |  |  | Venous |  |  |  |  |
| --- | --- | --- | --- | --- | --- | --- | --- | --- | --- | --- | --- |
|  |  | N | Sensitivity<br>(95% CI) | Specificity<br>(95% CI) | PPV<br>(95% CI) | NPV<br>(95% CI) | N | Sensitivity<br>(95% CI) | Specificity<br>(95% CI) | PPV<br>(95% CI) | NPV<br>(95% CI) |
| SD BIOLINE<br>Ag Pf<br>(#05FK50) | PfHRP2 | 220 | 0.714<br>(0.636 – 0.784) | 0.939<br>(0.852 – 0.983) | 0.965<br>(0.913 – 0.990) | 0.585<br>(0.485 – 0.680) | N/A | N/A | N/A | N/A | N/A |
| BIOCREDIT<br>Pf<br>(pLDH/HRPII) | Pf (HRP2<br>and/or PfLDH<br>positive) | 218 | 0.783<br>(0.709 – 0.846) | 0.894<br>(0.794 – 0.956) | 0.944<br>(0.889 – 0.977) | 0.641<br>(0.535 – 0.739) | 220 | 0.818<br>(0.748 – 0.876) | 0.894<br>(0.794 – 0.956) | 0.947<br>(0.895 – 0.979) | 0.678<br>(0.569 – 0.774) |
|  | PfHRP2 Only | 218 | 0.776<br>(0.702 – 0.840) | 0.924<br>(0.832 – 0.975) | 0.959<br>(0.908 – 0.987) | 0.642<br>(0.537 – 0.738) | 220 | 0.812<br>(0.741 – 0.870) | 0.924<br>(0.832 – 0.975) | 0.962<br>(0.913 – 0.987) | 0.678<br>(0.571 – 0.772) |
|  | PfLDH Only | 218 | 0.618<br>(0.536 – 0.696) | 0.955<br>(0.873 – 0.991) | 0.969<br>(0.912 – 0.994) | 0.521<br>(0.428 – 0.612) | 220 | 0.662<br>(0.582 – 0.736) | 0.970<br>(0.895 – 0.996) | 0.981<br>(0.932 – 0.998) | 0.552<br>(0.457 – 0.644) |
| BIOCREDIT<br>Pf/Pv<br>(pLDH/pLDH) | PfLDH | 218 | 0.645<br>(0.563 – 0.721) | 0.955<br>(0.873 – 0.991) | 0.970<br>(0.916 – 0.994) | 0.538<br>(0.444 – 0.631) | 220 | 0.662<br>(0.582 – 0.736) | 0.970<br>(0.895 – 0.996) | 0.981<br>(0.932 – 0.998) | 0.552<br>(0.457 – 0.644) |
| BIOCREDIT<br>Pf (pLDH) | PfLDH | 217 | 0.642<br>(0.560 – 0.719) | 0.955<br>(0.873 – 0.991) | 0.970<br>(0.915 – 0.994) | 0.538<br>(0.444 – 0.631) | 220 | 0.662<br>(0.582 – 0.736) | 0.970<br>(0.895 – 0.996) | 0.981<br>(0.932 – 0.998) | 0.552<br>(0.457 – 0.644) |
| Microscopy | Parasites | N/A | N/A | N/A | N/A | N/A | 183 | 0.534<br>(0.445 – 0.622) | 0.981<br>(0.897 – 1.000) | 0.986<br>(0.924 – 1.000) | 0.455<br>(0.361 – 0.552) |

*Abbreviations:* Ag, antigen; CI, confidence interval; HRP2, histidine-rich protein 2; *Pf*, *Plasmodium falciparum*; PfLDH, *Plasmodium falciparum*- specific lactate dehydrogenase; pLDH, *Plasmodium* lactate dehydrogenase; PPV, positive predictive value; *Pv*, *Plasmodium vivax*; NPV, negative predictive value; N/A, not applicable

### Diagnostic performance of investigational and comparator RDTs against antigen concentration

Sixty-six percent (131/200) of participants whose samples were tested with the quantitative antigen assay were *Pf*-positive. Table 3 presents a summary of the diagnostic performance of all investigational and comparator tests, compared to the quantitative antigen assay, by specimen type. When RDT performance was compared against the quantitative antigen assay as the reference method, the sensitivity of all tests on both capillary and venous specimens was significantly higher than when compared against the PCR reference, with the largest improvements observed on the PfLDH test lines. The same trend is observed for specificity, with the exception of the HRP2 line on the BIOCREDIT Malaria Ag Pf (pLDH/HRPII) test with capillary specimens, which showed 92% specificity (95% CI: 82.5 – 96.8), as compared to 92% specificity (95% CI: 82.5–96.8), compared to 92% (95% CI: 83.2–97.5) when compared against PCR. Overall, the PfLDH-only tests had the highest performance compared to the antigen assay, irrespective of specimen type. This can be understood in the context of the performance of the quantitative antigen assay relative to the reference PCR method, which also showed the lowest sensitivity (71%) for the detection of PfLDH (Supplementary Table 1). All RDTs performed slightly better on venous blood specimens than on capillary specimens using both reference methods, although differences were negligible and within the 95% CIs.

**Table 3.** Diagnostic performance of investigational and comparator tests against the reference quantitative antigen assay for detection of *P. falciparum* by specimen type.

| Test | Target | Capillary |  |  |  |  | Venous |  |  |  |  |
| --- | --- | --- | --- | --- | --- | --- | --- | --- | --- | --- | --- |
|  |  | *N* | *Sensitivity*<br>(95% *CI*) | *Specificity*<br>(95% *CI*) | *PPV*<br>(95% *CI*) | *NPV*<br>(95% *CI*) | *N* | *Sensitivity*<br>(95% *CI*) | *Specificity*<br>(95% *CI*) | *PPV*<br>(95% *CI*) | *NPV*<br>(95% *CI*) |
| SD BIOLINE<br>Ag Pf<br>(#05FK50) | PfHRP2 | 200 | 0.760<br>(0.677 – 0.831) | 0.958<br>(0.881 – 0.991) | 0.970<br>(0.916 – 0.994) | 0.687<br>(0.586 – 0.776) | N/A | N/A | N/A | N/A | N/A |
| BIOCREDIT<br>Pf<br>(pLDH/HRPII) | Pf (HRP2<br>and/or PfLDH<br>positive) | 198 | 0.837<br>(0.762 – 0.896) | 0.928<br>(0.839 – 0.976) | 0.956<br>(0.900 – 0.985) | 0.753<br>(0.647 – 0.840) | 200 | 0.893<br>(0.827 – 0.940) | 0.971<br>(0.899 – 0.996) | 0.983<br>(0.941 – 0.998) | 0.827<br>(0.727 – 0.902) |
|  | PfHRP2 Only | 198 | 0.819<br>(0.741 – 0.882) | 0.915<br>(0.825 – 0.968) | 0.945<br>(0.885 – 0.980) | 0.739<br>(0.634 – 0.827) | 200 | 0.876<br>(0.806 – 0.927) | 0.958<br>(0.881 – 0.991) | 0.974<br>(0.926 – 0.995) | 0.810<br>(0.709 – 0.887) |
|  | PfLDH Only | 198 | 0.869<br>(0.786 – 0.928) | 0.990<br>(0.945 – 1.000) | 0.989<br>(0.938 – 1.000) | 0.883<br>(0.808 – 0.936) | 200 | 0.911<br>(0.838 – 0.958) | 0.990<br>(0.945 – 1.000) | 0.989<br>(0.942 – 1.000) | 0.916<br>(0.846 – 0.961) |
| BIOCREDIT<br>Pf/Pv<br>(pLDH/pLDH) | PfLDH | 198 | 0.889<br>(0.810 – 0.943) | 0.980<br>(0.929 – 0.998) | 0.978<br>(0.922 – 0.997) | 0.898<br>(0.825 – 0.948) | 200 | 0.911<br>(0.838 – 0.958) | 1.000<br>(0.963 – 1.000) | 1.000<br>(0.961 – 1.000) | 0.917<br>(0.848 – 0.961) |
| BIOCREDIT<br>Pf (pLDH) | PfLDH | 197 | 0.898<br>(0.820 – 0.950) | 0.990<br>(0.945 – 1.000) | 0.989<br>(0.939 – 1.000) | 0.907<br>(0.836 – 0.955) | 200 | 0.911<br>(0.838 – 0.958) | 1.000<br>(0.963 – 1.000) | 1.000<br>(0.961 – 1.000) | 0.917<br>(0.848 – 0.961) |

*Abbreviations:* Ag, antigen; CI, confidence interval; HRPII/2, histidine-rich protein 2; *Pf*, *Plasmodium falciparum*; PfLDH, *Plasmodium falciparum*- specific lactate dehydrogenase; pLDH, *Plasmodium* lactate dehydrogenase; PPV, positive predictive value; *Pv*, *Plasmodium vivax*; NPV, negative predictive value; N/A, not applicable

### Antigen concentration distribution and performance of the BIOCREDIT Malaria Ag (pLDH/HRPII) RDT

Figure 1 summarizes the relationship between (A) HRP2 concentration and parasite density and (B) LDH concentration and parasite density, with color coding used to distinguish results for each line on the BIOCREDIT Malaria Ag Pf (pLDH/HRPII). The HRP2 line detects specimens with lower target antigen concentrations compared to the PfLDH line.

**Figure 1.**
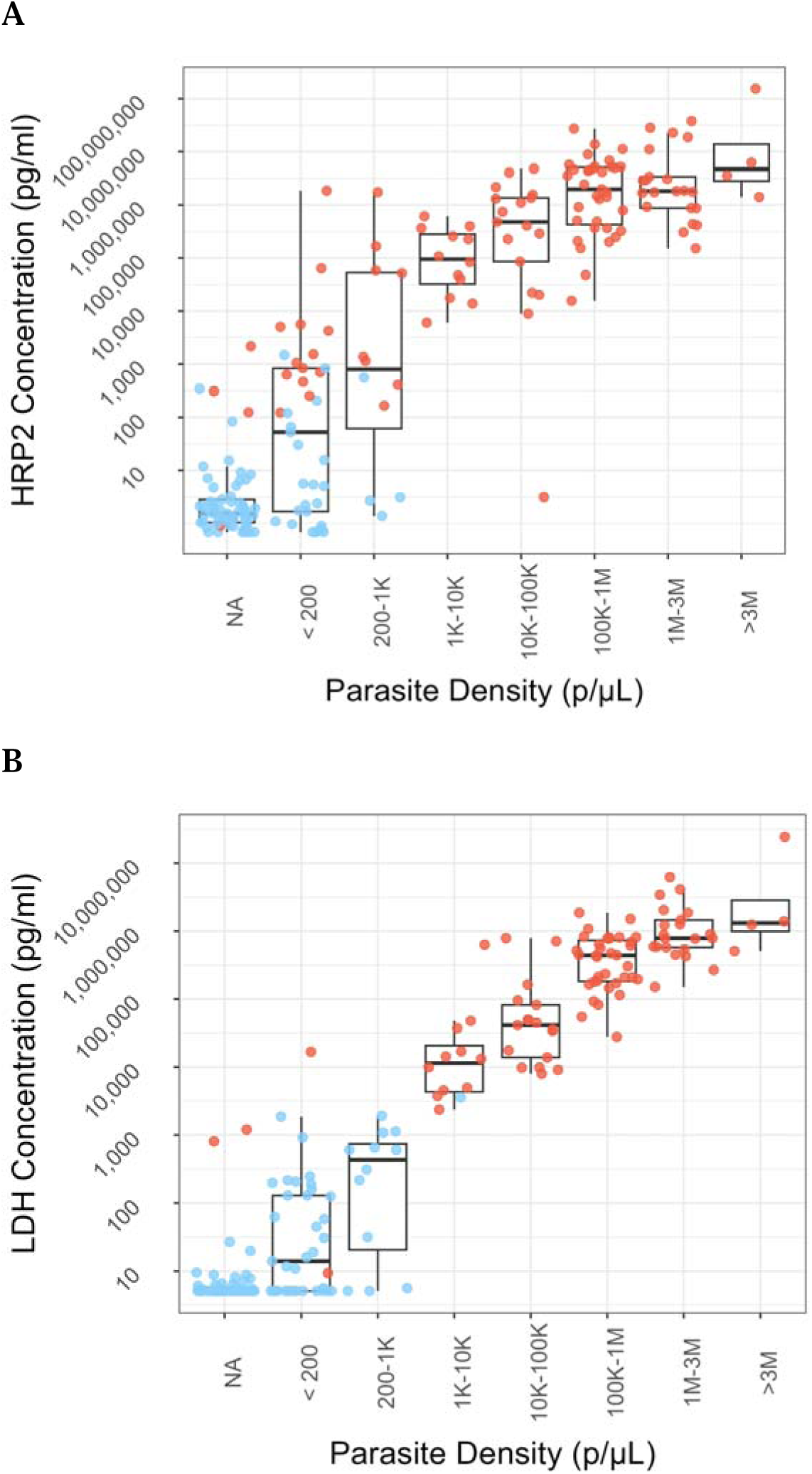
Box plots of (A) HRP2 and (B) pfLDH antigen concentration distributions as a function of parasite density in PCR-confirmed cases. Results by line from the BIOCRDIT Pf (pLDH/HRPII) test are indicated in color. Positive test lines are shown in red, and negative test lines are shown in blue for the HRP2 line and pfLDH line in panels A and B, respectively.

The BIOCREDIT Malaria Ag Pf (pLDH/HRPII) RDT has a 50% probability of detecting HRP2 at an antigen concentration of 0.175 ng/mL, and a 90% probability at 2.84 ng/mL (Supplementary Figure 1). The 50% and 90% probabilities of detection for PfLDH are 2.17 ng/mL and 12.24 ng/mL, respectively (Supplementary Figure 1).

Figure 2 shows the contribution of each test line to the overall performance of the BIOCREDIT Malaria Ag Pf (pLDH/HRPII), displaying the PfLDH and HRP2 concentrations for each clinical specimen. The results for the HRP2 and PfLDH test lines on the RDT are shown for each clinical specimen in panels A and B, respectively, and combined in panel C. More PCR-confirmed specimens had detectable HRP2 concentrations on the HRP2 line but not the LDH line, compared to specimens that were detectable on the LDH line but not the HRP2 line (Figure 2, panel C).

**Figure 2.**
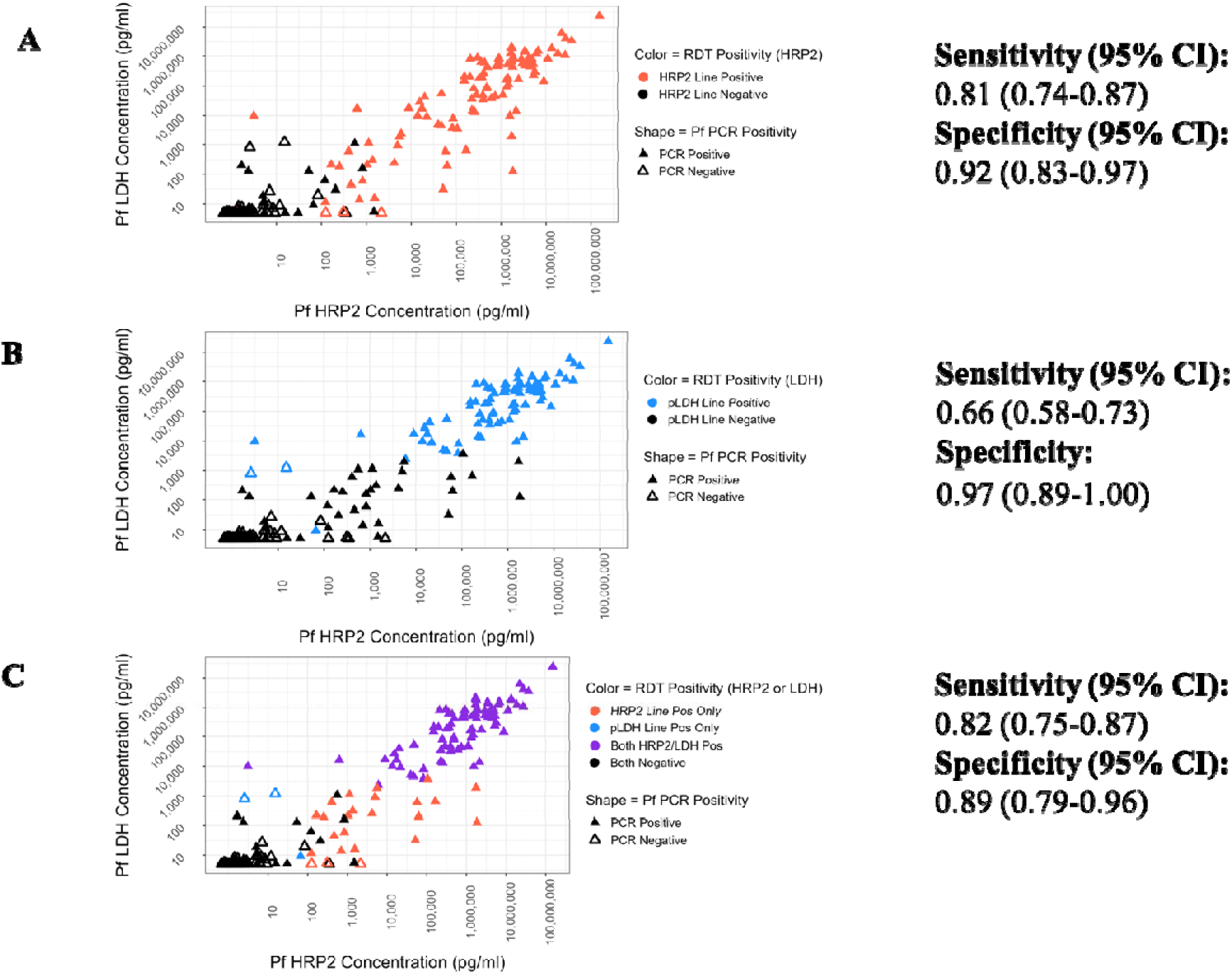
HRP2 concentration plotted against PfLDH for clinical specimens. PCR- confirmed *P. falciparum* specimens are shown as filled triangles. PCR-negative specimens are represented as an open triangle. Test line results from the BIOCREDIT Pf (pLDH/HRPII) test are represented in color: red for HRP2-positive specimens, blue for LDH-positive specimens, and purple for specimens positive on both lines (Panel C).

RDT line intensity was also correlated with the antigen concentration for the BIOCREDIT Pf (pLDH/HRPII) test (Figure 3). The HRP2 line showed an antigen-dependent increase in visible intensity between approximately 0.1 ng/mL and 100 ng/mL (100 –100,000 pg/mL). The PfLDH line showed an antigen-dependent increase in visible intensity between approximately 2 ng/mL and 100 ng/mL (2,000 – 100,000 pg/mL). For both lines, the test lines for specimens with HRP2 or PfLDH concentrations above 100 ng/mL appeared to be saturated and did not significantly increase with higher antigen concentrations.

**Figure 3.**
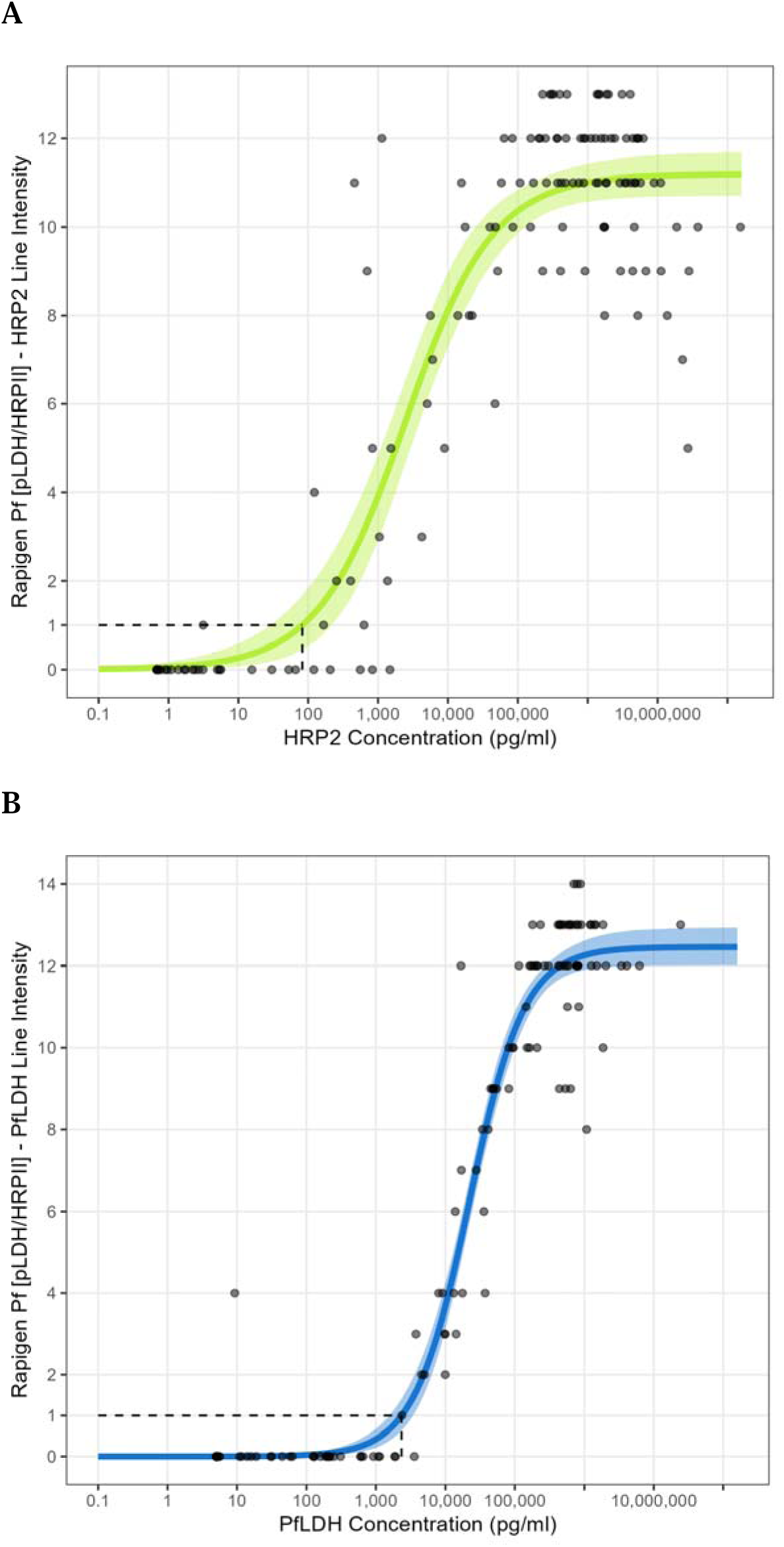
Correlation between antigen concentration and line intensity (0 is a negative test result, 1-15 are positive test results) on the BIOCREDIT Pf (pLDH/HRPII) RDT. Panel A shows the intensity of the HRP2 line on the RDT plotted against the HRP2 concentration. Panel B shows the intensity of the LDH line on the RDT plotted against LDH concentration. Dotted lines indicate the concentration at which line intensity is weakly visible (score of 1).

### Performance prediction based on analytical sensitivity

Table 4 shows the predicted sensitivities based on the test’s analytical LOD as applied to this study population against both the quantitative antigen and PCR reference methods and the observed sensitivities (also shown in Tables 2 and 3) for each antigen on the RDT and the combined RDT test result. The specificity against the quantitative antigen reference method is, by definition, 100% for all antigens; therefore, only specificity against PCR is presented. Overall, the predicted and observed sensitivities were comparable. When using the antigen concentration reference method, the predicted sensitivity of the RDT was slightly lower than the observed sensitivity for the detection of HRP2 (84% versus 88%) but slightly higher for the detection of PfLDH (92% versus 91%). For the detection of either HRP2 and/or PfLDH, the predicted sensitivity was lower than the observed sensitivity (84% versus 89%). Against the PCR reference method, the test’s predicted sensitivity was lower than the observed sensitivity for HRP2 (78% versus 81%) and slightly higher for PfLDH (67% versus 66%). For HRP2 and/or PfLDH, the predicted sensitivity was, again, lower (78% versus 82%).

**Table 4.** Performance of the Rapigen BIOCREDIT Pf (pLDH/HRPII) RDT. Sensitivity is presented against confirmed positive cases separately by the quantitative antigen and PCR reference assays. Specificity is presented against confirmed negative cases by PCR. The table presents (i) the predicted results of the test based on its analytical limit of detection, and (ii) the observed results of the test against each reference method.

| Test Line | Quantitative antigen assay reference |  | PCR reference |  |  |  |
| --- | --- | --- | --- | --- | --- | --- |
|  | Sensitivity (95% CI) |  | Sensitivity (95% CI) |  | Specificity (95% CI) |  |
|  | Predicted | Observed (Table 3) | Predicted | Observed (Table 2) | Predicted | Observed (Table 2) |
| HRP2 line | 0.844<br>(0.769 – 0.902) | 0.876<br>(0.806 – 0.927) | 0.775<br>(0.697 – 0.842) | 0.812<br>(0.741 – 0.870) | 0.984<br>(0.912 – 1.000) | 0.958<br>(0.881 – 0.991) |
| PfLDH line | 0.920<br>(0.848 – 0.965) | 0.911<br>(0.838 – 0.958) | 0.667<br>(0.581 – 0.745) | 0.662<br>(0.582 – 0.736) | 1.000<br>(0.941 – 1.000) | 0.990<br>(0.945 – 1.000) |
| HRP2 and/or PfLDH lines | 0.838<br>(0.764 – 0.897) | 0.893<br>(0.827 – 0.940) | 0.783<br>(0.704 – 0.848) | 0.818<br>(0.748 – 0.876) | 0.984<br>(0.912 – 1.000) | 0.971<br>(0.899 – 0.996) |

Abbreviations: CI, confidence interval; HRP2, histidine-rich protein 2; PCR, polymerase chain reaction; Pf, *Plasmodium falciparum*; PfLDH, *Plasmodium falciparum*-specific lactate dehydrogenase; pLDH, *Plasmodium* lactate dehydrogenase; PPV, positive predictive value; NPV, negative predictive value; N/A, not applicable

For overall test performance against PCR, the HRP2 line was predicted to contribute the most to test sensitivity, with a marginal increase when the LDH line was included. Indeed, this pattern of performance was observed in the study. Additionally, the HRP2 line was predicted to contribute to a drop in the specificity of the test compared to LDH line when using the PCR reference method, as was observed in the study.

### Usability results

In total, 10 healthcare workers evaluated the usability of the BIOCREDIT Pf (pLDH) test, and 16 evaluated the BIOCREDIT Pf (pLDH/HRPII) test. All participants successfully conducted the tests, and no critical errors were observed during the use of either test. Responses to the multiple-choice label comprehension questionnaires indicated that most participants understood the tests’ intended uses, safety information, and warnings (Supplementary Table 2). The greatest source of error in the test’s workflow was in identifying the correct number of assay buffer drops to add to the test devices and determining the correct reading window for test result interpretation. Additionally, most contrived test result images were correctly read and interpreted by participants, with an overall interpretation error rate of 4.0% for the BIOCREDIT Pf (pLDH) test and 4.8% for the BIOCREDIT Pf (pLDH/HRPII) test (Supplementary Table 3). Ninety percent of participants evaluating the BIOCREDIT Pf (pLDH) test reported that it was either “easy” or “very easy” to use, compared to 70% of those evaluating the BIOCREDIT Pf (pLDH/HRPII) test.

## Discussion

This study evaluated the performance of three novel LDH-based malaria RDTs in a febrile population in Kédougou, Senegal, and assessed RDT performance as a function of antigen concentration distribution. Improved sensitivity for PfLDH is critical for RDTs to detect emerging *hrp2/3* deletions. The WHO recommends switching to LDH-based RDTs only when there is a confirmed significant prevalence (>5%) of false-negative RDT results arising from *hrp2/hrp3* deletions due to the documented lower sensitivity of LDH-based RDTs for *Pf*.^16^ This study population had only one specimen with a suspected deletion based on the HRP2 and PfLDH results from the antigen quantification assay; however, this case could also be attributed to a *Plasmodium malariae* or *ovale* infection, which have previously been found in this area of Senegal.^27^ Specimens in this study were not genotyped for *hrp2/3* deletions.

Among all RDTs included in this study, the BIOCREDIT Malaria Ag Pf (pLDH/HRPII) test, which has test lines for both HRP2 and PfLDH, showed the highest sensitivity for the detection of *Pf* at 78%. This sensitivity is slightly higher than that of the SD Bioline HRP2-based RDT (71%), an established, WHO-prequalified product currently in routine use in Senegal. Although the HRP2 line alone on the BIOCREDIT Malaria Ag Pf (pLDH/HRPII) was more sensitive than the comparator SD Bioline HRP2- based test (78% versus 71% on capillary specimens), the sensitivities of the PfLDH lines alone on all three BIOCREDIT RDTs (61%–64% on capillary specimens) were lower than that of the comparator.

This suggests that in populations such as this, where *hrp2/3* deletions are uncommon, the PfLDH line alone cannot compensate for the performance of the HRP2 line, even with the improved LOD for LDH on the Rapigen tests.

On the BIOCREDIT Malaria Ag Pf (pLDH/HRPII) RDT, a higher rate of false positives was observed on both capillary and venous specimens when considering results based on either test line, leading to a consequently lower specificity than other RDTs included in this evaluation. For most of these specimens (5/7), the measured antigen concentrations were below the detection levels or lower than the LOD of the RDT. Some drop in specificity may be attributed to the persistence of HRP2, as shown in the predicted performance of the RDT compared to the PCR reference assay, in contrast to LDH, where the presence of LDH closely corresponds to the presence of parasite DNA.

The results of this study are consistent with findings from other clinical evaluations of these tests. In Burundi, Niyukuri *et al.* (2022) reported sensitivities of 79.9% and 72.3% for the BIOCREDIT Malaria Ag Pf (pLDH/HRPII) test in clinical and subclinical populations, respectively, in a sample with only two *hrp3* deletions.^21^ The authors also similarly reported lower specificity for this test compared to the HRP2- only comparator RDT in both clinical and subclinical cases (82.4% versus 96.2% for clinical cases; 84.4% versus 93.4% for subclinical cases). Another study conducted in Djibouti, where there are high rates of *hrp2/3* deletions and *Pv* cases,^33–35^ found a sensitivity and specificity of 88.2% and 100%, respectively, on the BIOCREDIT Malaria Ag Pf/Pv (pLDH/pLDH) for the detection of *Pf.*^23^

The inclusion of a quantitative assay to determine antigen concentration in this study allowed for assessment of RDT performance relative to the cognate analytes detected by each RDT test line. The performance of the RDT, evaluated against a reference assay for the same analyte (in contrast to microscopy or PCR), demonstrated improved sensitivity and specificity for both antigens across all RDTs. The relative sensitivities of the individual antigen test lines, when using the quantitative antigen reference assay compared to PCR/DNA as a reference standard, are driven by the analytical sensitivity of the reference assay for the analyte, the overall antigen concentration distribution in the specimens, and the absolute LODs of the RDTs for the antigens. The quantitative antigen assay has a higher clinical LOD for LDH compared to HRP2,^29^ which results in missing clinical samples with low LDH concentrations.

Additionally, clinical specimens overall have lower LDH concentrations compared to HRP2, resulting in fewer PCR samples with measurable LDH levels, as shown previously.^29,30^ Consequently, the sensitivity of the antigen lines on the RDTs is highest for LDH when using the antigen reference method but lowest when using the PCR method. Most essentially, the lower analytical sensitivity of the LDH lines combined with the lower abundance of LDH in clinical specimens compared to HRP2 results in a lower sensitivity of the LDH lines compared to the HRP2 lines in the RDTs evaluated in this study.

Although not an intended endpoint of the study, the training of test operators to record RDT line intensity in this study enabled the demonstration that—even by eye—a reasonable relationship was present between RDT line intensity and antigen concentration. Given that challenge and longitudinal studies suggest that the ratio of HRP2 to LDH can be used to differentiate active from recently cleared infections,^36^ further investigation into the utility of HRP2/LDH line intensities from an RDT for this purpose is warranted.

The analytical sensitivity (LOD) of the BIOCREDIT Malaria Ag Pf (pLDH/HRPII) RDT was previously determined through laboratory-based benchmarking studies.^20^ In this study, the laboratory-derived LOD of the BIOCREDIT Malaria Ag Pf (pLDH/HRPII) test was combined with the distribution of antigen concentrations and reference PCR results from the study population to estimate the predicted performance of the RDT, which was then compared to the observed performance from this study. Overall, the predicted performance of the test was comparable to the observed performance, demonstrating the value of understanding the underlying antigen concentration in a population and validating the ability to predict RDT performance based on analytical LODs. Notably, as we observed, the predictions confirmed HRP2 as the largest driver of clinical sensitivity for the test in this population, with LDH contributing to a marginal improvement in clinical sensitivity for *Pf* infection. Additionally, as observed, there is a larger drop in specificity due to the HRP2 line in contrast to the LDH line. This predicted drop in specificity arises exclusively from specimens which have antigen but are PCR negative. The drops in specificity are driven either by the emergence of antigen in advance of DNA, or more likely, by the accumulation and persistence of antigen after DNA clearance. Other sources of cross-reactivity not accounted for in analytical studies are likely to contribute to the actual observed clinical specificity.

Using analytical performance to predict the performance of new RDTs in different populations with varying parasite density distributions is a valuable tool for assessing the impact of new tests in diverse contexts of use. As previously shown for SARS-CoV-2 RDTs, where quantitative N-antigen data could be used to predict clinical sensitivity across different specimen types,^37,38^ understanding the underlying HRP2 and LDH concentration distributions across various use cases and epidemiological settings (e.g., case management, asymptomatic, pregnancy, low and high transmission) can improve predictions of test performance in these contexts.

From a usability perspective, the study demonstrated that intended users in this setting were able to accurately comprehend key elements of the product labels and correctly interpret images of results. This aligns with a long history of prior research demonstrating the simplicity of these tests and their ability to be conducted by end users.^39–41^ The most common errors related to the correct identification of the number of assay buffer drops to add to the test devices and determining the appropriate reading window for interpreting test results. These aspects of malaria RDT workflows often vary between products, highlighting the importance of context-specific training that emphasizes key aspects of the test procedure.^42^ The results suggest a slight user preference for the *Pf* RDT with a single test line compared to the product with two separate test lines for HRP2 and PfLDH. However, both products were evaluated by small samples and two different groups of users; therefore, comparisons should be interpreted with caution.

Several other limitations of this study should also be noted. Microscopy and antigen quantification results were only available for a subset of participants. Although *Pv* has been observed in Eastern Senegal,^13,27,43,44^ no cases were identified in this population. Consequently, the performance of the BIOCREDIT Malaria Ag Pf/Pv (pLDH/pLDH) test for the detection of *Pv* malaria was not assessed, and test performance on specimens with mixed infections could not be evaluated. Lastly, this study assessed test performance in a symptomatic, febrile population in an area of high endemicity. Analytical benchmarking and prior evaluations suggest that relative performance improvements may be expected with asymptomatic populations and subclinical cases.^20,21,24^ Future studies should further investigate test performance in these contexts.

In summary, this study confirms that despite the higher analytical sensitivity of the BIOCREDIT tests’ LDH line compared to other currently WHO-prequalified RDTs, the HRP2 line primarily drives the sensitivity of the RDT in this high-burden setting with negligible suspected *hrp2/hrp3* deletions. An RDT that performs equally to support clinical diagnosis of *Pf* malaria, regardless of the underlying prevalence of *hrp2/hrp3* deletions, should detect both HRP2 and LDH, possibly on the same line to simplify interpretation.

## Data Availability

The data underlying the results presented in this manuscript can be accessed at the following link: https://doi.org/10.7910/DVN/0OMVWZ

## Acknowledgments

The authors appreciate the support and contributions of the study participants. We also thank the research staff and laboratory technicians at the study sites and the Institut Pasteur de Dakar for their valuable support. We thank Mikka Nyarko for editorial support provided to this manuscript.

## Conflict of interest statement

The authors have no competing interests to declare.

## Funding statement

This work was supported by grants from the Bill & Melinda Gates Foundation (INV-006979 and INV- 071824) to GD. Rapigen donated the BIOCREDIT RDTs used in this evaluation. The REDCap instance used is supported by the Institute of Translational Health Sciences, which is funded by the National Center for Advancing Translational Sciences of the National Institutes of Health under award number UL1TR002319.

---

**Supplementary Figure 1.**
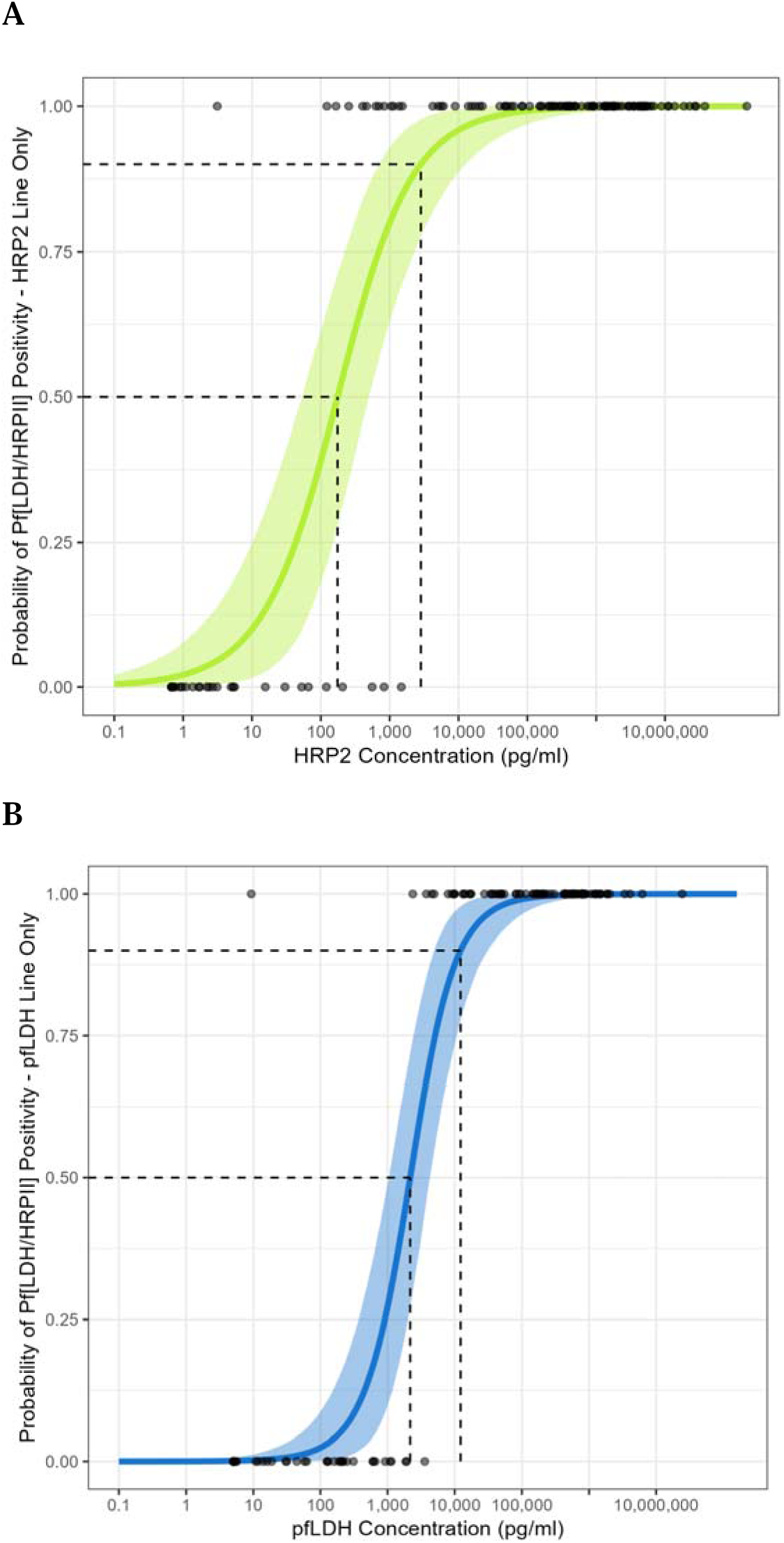
Probability of test line positivity on the BIOCREDIT Pf (pLDH/HRPII) RDT as a function of antigen concentration. In panel A, the probability of the HRP2 test line positivity is plotted against HRP2 concentration. In panel B, the probability of the LDH test line positivity is plotted against LDH concentration. The 50% and 90% probabilities of positivity are shown on both graphs.

**Supplementary Table 1.** Diagnostic performance of the quantitative antigen concentration assay against the reference PCR for the detection of *P. falciparum*.

| Test | Target | Venous |  |  |  |  |
| --- | --- | --- | --- | --- | --- | --- |
|  |  | *N* | *Sensitivity*<br>(95% *CI*) | *Specificity*<br>(95% *CI*) | *PPV*<br>(95% *CI*) | *NPV*<br>(95% *CI*) |
| Quantitative antigen assay | PfHRP2 | 199 | 0.855<br>(0.785 – 0.909) | 0.836<br>(0.719 – 0.918) | 0.922<br>(0.861 – 0.962) | 0.718<br>(0.599 – 0.819) |
|  | PfLDH | 199 | 0.710<br>(0.627 – 0.784) | 0.967<br>(0.887 – 0.996) | 0.980<br>(0.930 – 0.998) | 0.596<br>(0.493 – 0.693) |
|  | Pf (HRP2 and/or PfLDH positive) | 199 | 0.862<br>(0.793 – 0.915) | 0.820<br>(0.700 – 0.906) | 0.915<br>(0.854 – 0.957) | 0.725<br>(0.604 – 0.825) |

Abbreviations: CI, confidence interval; HRP2, histidine-rich protein 2; PCR, polymerase chain reaction; PfLDH, *Plasmodium falciparum*-specific lactate dehydrogenase; PPV, positive predictive value; NPV, negative predictive value

**Supplementary Table 2.**
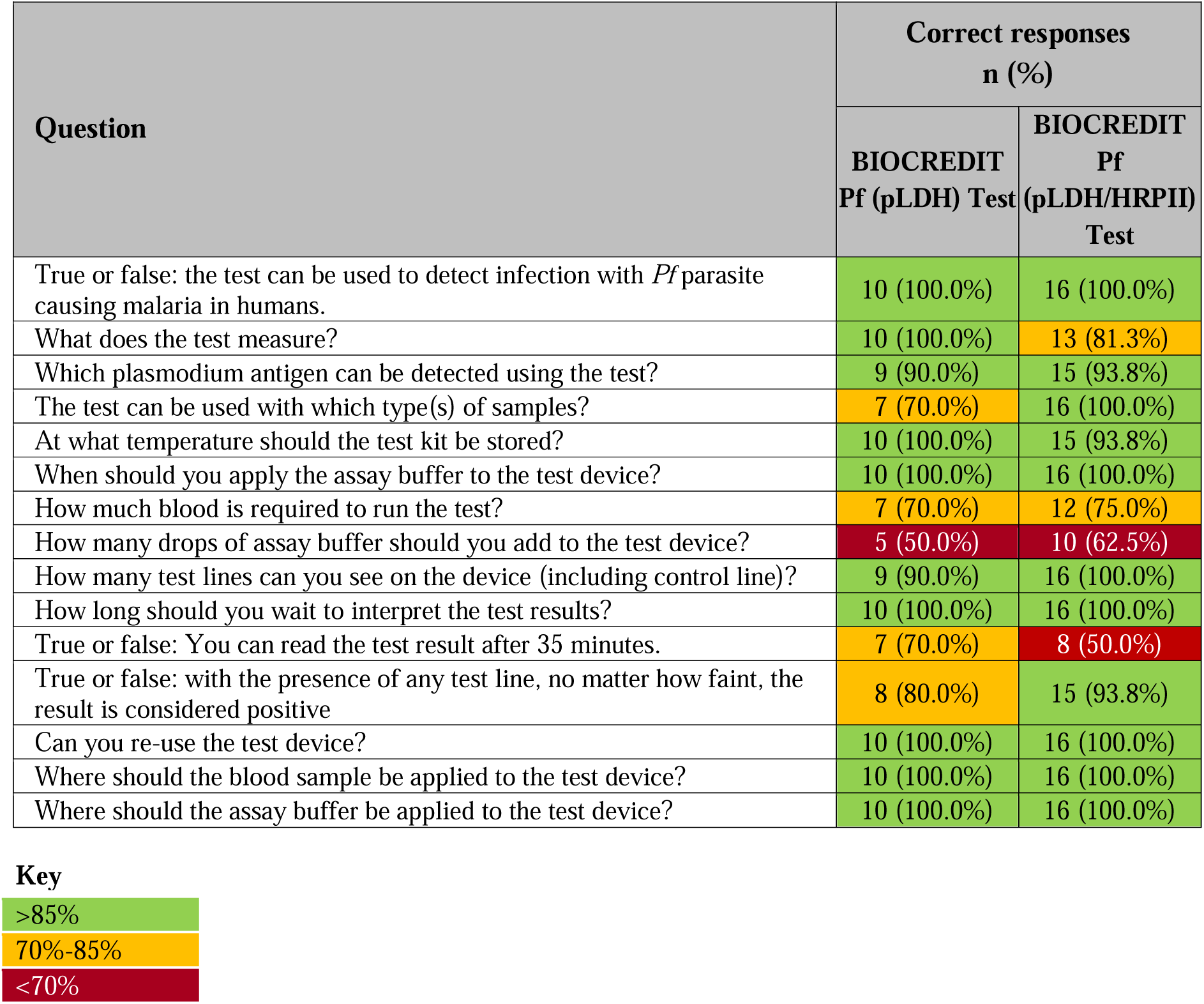
Label comprehension questionnaire results for the Pf (pLDH) and Pf (pLDH/HRPII) tests.

**Supplementary Table 3.**
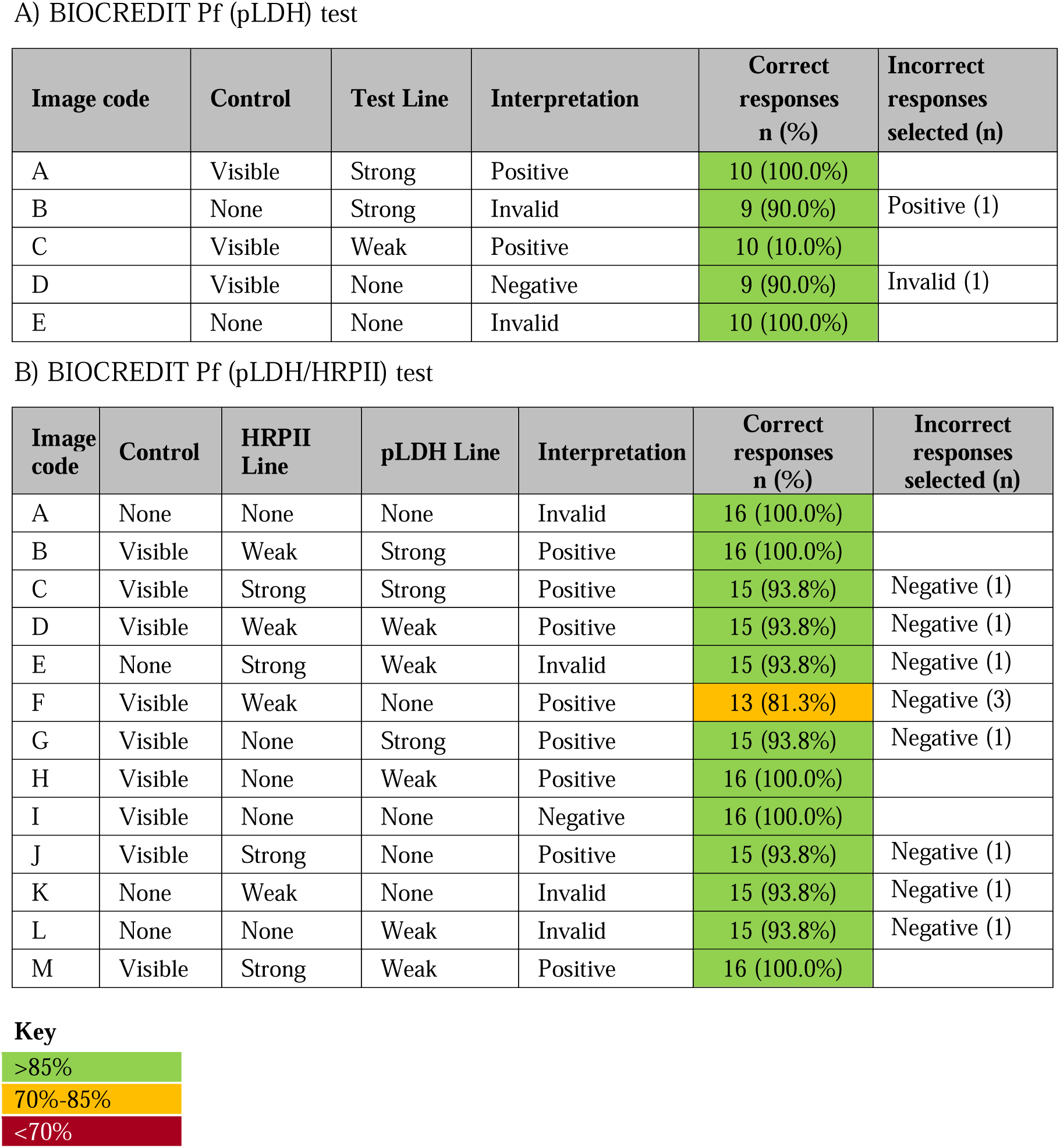
Result interpretation questionnaire results for (a) the Pf (pLDH) test, and (b) the Pf (pLDH/HRPII) test.

## Notes

### Competing Interest Statement

The authors have declared no competing interest.

